# High levels of IsoDGR-modified fibronectin are associated with higher levels of macrophages and other rupture-prone plaque characteristics in patients undergoing carotid endarterectomy

**DOI:** 10.1101/2024.09.02.24312942

**Authors:** Tetiana Motsak, Barend M. Mol, Joost K.R. Hoekstra, Valentina Osorio Zuluaga, Saskia Rumpff-Derksen, Ioanna Paspali-Strik, Sofong Cam Ngan, Gerard Pasterkamp, Gert J. de Borst, Siu Kwan Sze, Dominique P.V. de Kleijn

## Abstract

**Objective:** Deamidation of the NGR (Asn-Gly-Arg) motif to the isoDGR (isoAsp-Gly-Arg) motif in fibronectin (IsoDGR-fibronectin) enhances in vitro monocyte and endothelial cell activation. Blocking isoDGR reduces macrophage influx in murine tissues. Although macrophage influx is an important feature of human plaque destabilization, the role for plasma and plaque isoDGR-fibronectin in macrophage influx in the atherosclerotic plaque and thereby increasing plaque vulnerability has not been investigated in large human cohorts.

**Design:** IsoDGR-fibronectin levels in plasma and plaques were measured in carotid endarterectomy (CEA) patients from the Athero Express biobank cohort and associated with macrophage and other vulnerable plaque characteristics in the carotid plaque of the same patient.

**Methods:** Levels of isoDGR-fibronectin were measured using an ELISA. Carotid plaque characteristics were visualized with immunohistochemistry staining and scored semi-quantitatively. Baseline characteristics were analysed with Pearson’s Chi-squared test and Mann-Whitney U-test when applicable. Univariate and multivariate logistics regression analyses were used to identify associations with adverse plaque characteristics.

**Results:** Plasma isoDGR-fibronectin was measured in 730 CEA patients. Patients with moderate/heavy plaque macrophage staining had higher levels of isoDGR-fibronectin than patients with no/minor macrophage staining (multivariate OR 1.40 (95%CI 1.04 – 1.90, p=0.028)). Of the 730 CEA patients, 348 had plaque samples available for isoDGR-fibronectin measurements. In the multivariate analysis, higher plaque levels of isoDGR-fibronectin were associated with moderate/high plaque macrophage staining (OR 1.22 (95%CI 1.00 – 1.56, p=0.049)), >40% fat in plaque (OR 1.1.44 (95% CI 1.14 – 1.86, p=0.004)) and intraplaque haemorrhage (OR 1.38 (95% 1.12 – 1.72, p=0.003)).

**Conclusion:** In this large human cohort study high plasma and plaque levels of isoDGR-fibronectin were associated with more plaque macrophages and other adverse plaque characteristics. This suggests the involvement of isoDGR-fibronectin in human plaque destabilization that may lead to new potential treatment modalities.

## Introduction

Atherosclerotic cardiovascular disease (ASCVD) is a highly prevalent worldwide disease with estimations of 500 million affected people and contributing to eighteen million annual deaths.^1,2^ Atherosclerosis has been considered for decades as an inflammatory disease^3^, but only recent clinical trials like CANTOS^4^ and LoDoCo2^5^ showed that inhibition of inflammation can reduce cardiovascular event like a stroke or myocardial infarction. Inflammation in atherosclerotic plaques develop over decades, leading to complex lesions with a thin fibrous cap and high influx of macrophages. These features can cause plaque rupture or plaque erosion, resulting in cardiovascular events.^6^ Other vulnerable plaque characteristics comprise a large lipid core, intraplaque hemorrhage and low levels of smooth muscle cells.^7–9^

Fibronectin is an adhesion molecule ubiquitously expressed in the extracellular matrix of cells mediating in numerous functions such as inflammation and hemostasis.^10^ Fibronectin and its splice variants are present in atherosclerotic fibrous cap formation but not in healthy arteries.^11^ Recently, we showed that deamidation of fibronectin activates monocytic and endothelial cells to promote atherosclerosis.^12^

In the process of deamidation, a spontaneous intramolecular rearrangement occurs in which a functional amide group in an amino acid such as asparagine (Asn) is converted into either aspartic acid (Asp) or iso-aspartic acid (isoAsp).^13^ When deamidation occurs in the NGR (Asn-Gly-Arg) motif in fibronectin into the isoDGR (isoAsp-Gly-Arg) motif, it increases monocyte adhesion in vitro^12,14,15^, suggesting that protein deamidation may promote human atherosclerosis by enhancing monocyte recruitment. This was confirmed in Pcmt1 ^-/-^ mice. Pcmt1 ^-/-^ mice cannot repair deamidation and accumulate isoDGR motifs, but when treated with a monoclonal antibody targeting isoDGR, macrophage influx in body tissues are reduced significantly.^16^

For this, we hypothesize that high levels of isoDGR-fibronectin in human plasma and plaques are associated with high numbers of macrophages in the human atherosclerotic carotid plaque. We used the Athero Express biobank in which plasma, plaque and clinical data are collected from carotid stenosis patients after carotid endarterectomy. Although in vitro and mouse studies strongly suggest a role of isoDGR-fibronectin in atherosclerotic plaque progression and specifically the influx of macrophages, this has not been investigated in large human cohorts.

Our study is the first study to investigate the role of isoDGR-fibronectin levels in plasma and atherosclerotic plaques and its association with macrophage staining in the plaque and other adverse plaque characteristics. This will support a role for isoDGR-fibronectin in human plaque progression and identify isoDGR as a potential target to intervene in atherosclerosis.

## Methods

### Study population

This study was done using plasma and plaque samples of carotid endarterectomy patients from the Athero Express biobank study with available citrate samples and full three years of follow-up. An in-depth study design of the Athero Express has been published earlier.^17^ In short, patients were included in two tertiary referral hospitals (University Medical Center Utrecht and St Antonius Hospital Nieuwegein, the Netherlands) from 2002 onwards. The day before surgery, informed consent is taken and various venous blood samples are collected, centrifuged and its plasma stored at -80 °C. The atherosclerotic plaque is harvested during endarterectomy surgery, divided into 5 mm segments and processed following standard procedure. The culprit lesion is paraffin embedded of which 5 µm thick sections are cut for histological staining. All other plaque segments are crushed in liquid nitrogen for protein and RNA isolation using 1 ml TripPure Isolation Reagent. Baseline characteristics and three-year follow-up is obtained with questionnaires and verified with medical records. The Athero Express biobank is approved by the Ethical Review Board of both institutions and conducted according to the declaration of Helsinki.^18^

### Immuno-histology

For immune-histology on the culprit lesion, the following stainings were performed according to the Athero Express protocol^17^: Picro Sirius red for collagen staining, CD68 for macrophages, alfa actin for smooth muscle cells (SMCs), haematoxylin for assessment of thrombus, calcification and fat and CD34 for microvessels. All stainings were semi-quantitively scored and for this study dichotomised. Presence of macrophages, SMCs, calcification and collagen as no/minor or moderate/heavy, density of microvessels in the plaque was determined by taking the average number of vessels at three hotspots and this was dichotomized over the group median. For the assessment of lipid content, the percentage of lipid in the total plaque was measured and dichotomized in above and below 40%. Intraplaque haemorrhage was scored either as present or absent. A plaque vulnerability score was calculated by assigning either 0 or 1 point for each individual risk feature: moderate/heavy macrophage and calcification staining, low/moderate staining for SMC’s, presence of intraplaque haemorrhage, vessel density higher than median and a lipid core of >40%, as was published earlier.^7^

### Plaque protein isolation

Using standard Athero Express protocol for protein isolation, each segment of plaque material was ground in a mortar with a cold pestle using liquid nitrogen and put in a tube with zirconium oxide beads. For protein isolation, Tris (pH 7,5 40mM) with protease inhibitors was added to the samples before undergoing two cycles of bead beating to facilitate lysis. Afterwards, the samples were centrifuged for 5 minutes at 25.000x G at 4 °C. The supernatant was filtered using a Spin-X filter and centrifuged for 10 minutes at 16.000x G at 4 °C. Supernatant with all soluble proteins was collected, aliquoted, and stored at -80 °C.

### Enzyme-Linked ImmunoSorbent Assay (ELISA)

IsoDGR-fibronectin was measured in plasma and plaques of CEA patients with a double sandwich Enzyme-Linked Immunosorbent Assay (ELISA). For plasma, a Nunc Maxisorp plate was coated with 0.5 µg/ml isoDGR antibody (Ab) in 100 mM carbonate-bicarbonate buffer pH 9.4. Plates were incubated overnight at 4 °C. The following day, the Ab was discarded and the wells were washed with PBS. The plates were blocked with PBS-5% Skimmed Milk (PBS-M) for 1.5 hours at room temperature (RT). After blocking, the PBS-M was discarded, and the plates were washed with PBS 0.05% Tween20 (PBS-T). Next, the plates were incubated with citrate plasma of the CEA patients. This was done for two hours at RT on a plate shaker (150 rpm). The plasma was discarded, and the plates were washed with PBS-T. The plates were blocked with PBS-M for one hour at RT. The PBS-M was discarded, and the plates were washed with PBS-T. Next, 1 µg/ml of biotinylated anti-hFibronectin was added in PBS-M, and incubated for one hour at RT. The biotinylated anti-hFibronectin was discarded and the plates were washed with PBS-T. The plates were incubated with 1 ug/ml streptavidin-HRP Ab in PBS-M for one hour at RT. The plates were washed with PBS-T. Afterwards, TMB substrate was added and incubated in the dark for 30 minutes. Lastly, the reaction was stopped with a stop reagent provided by Sigma-Aldrich. The optical density (OD) values were corrected for background signal and measured at 450 nm. As a standard curve, citrate blood plasma samples from healthy anonymous blood volunteers from the blood bank of the UMC Utrecht were used in different dilutions and with PBS-M as a blank. For IsoDGR-fibronectin measurements in plaque tris lysates the same protocol was used with the exception of Tris buffer being used as a blank.

### Statistical analysis

Baseline characteristics were divided in no/minor and moderate/heavy macrophage staining groups and shown as mean (standard deviation [SD]) or median (interquartile range [IQR]) when applicable. Categorical variables were reported in absolute numbers and percentages. Independent sample t-tests, Mann–Whitney U-tests and Pearson’s Chi Square test were used when applicable. Differences in plaque levels of isoDGR-fibronectin in different histology staining groups was analysed using a Mann-Whitney U-test and presented in boxplots. Both univariate and multivariate logistic regressions analysis were used to analyse the association between plaque and plasma levels of isoDGR-fibronectin and adverse plaque characteristics. To comply with assumptions, ELISA results were transformed with the negative natural logarithm and multiplied by 10 000 for logistics regression analysis. The multivariate analysis was corrected for common cardiovascular confounders: age, sex, BMI, current smoking status, hypertension, diabetes, kidney function, statin or other lipid lowering medication use, pre-operative symptoms, history of coronary artery disease and history of peripheral arterial occlusive disease. The association between plasma and plaque levels of isoDGR-fibronectin and the plaque vulnerability score (range 0-7) was tested with a univariate and multivariate linear regression model. The multivariate models were corrected for the aforementioned confounders. Two-sided p-values of <.05 were considered statistically significant. All analyses were performed using RStudio version 4.2.1. (R Foundation, Vienna, Austria).

## Results

### Baseline characteristics

Plasma isoDGR-fibronectin was measured in a cohort of 730 CEA. Plaque macrophage staining was available in 645 of the 730 patients with no differences in patient characteristics between the complete cohort and patients with available macrophage staining.

Patients with moderate/heavy macrophage staining were more likely to be male (79% vs 66%, p<.001) and were less likely to have a history of peripheral artery occlusive disease (19% vs 27%, p=.015). Although not significant in the pre-hoc Pearson’s Chi-squared test, plaques of asymptomatic patients were more likely to show no/minor macrophage staining (16% vs 11%, p=.091) and plaques of patients with stroke prior to surgery were more likely to have plaques with moderate/heavy macrophage staining (27% vs 23%, p=.091). See table 1 for all baseline characteristics.

**Table 1.**
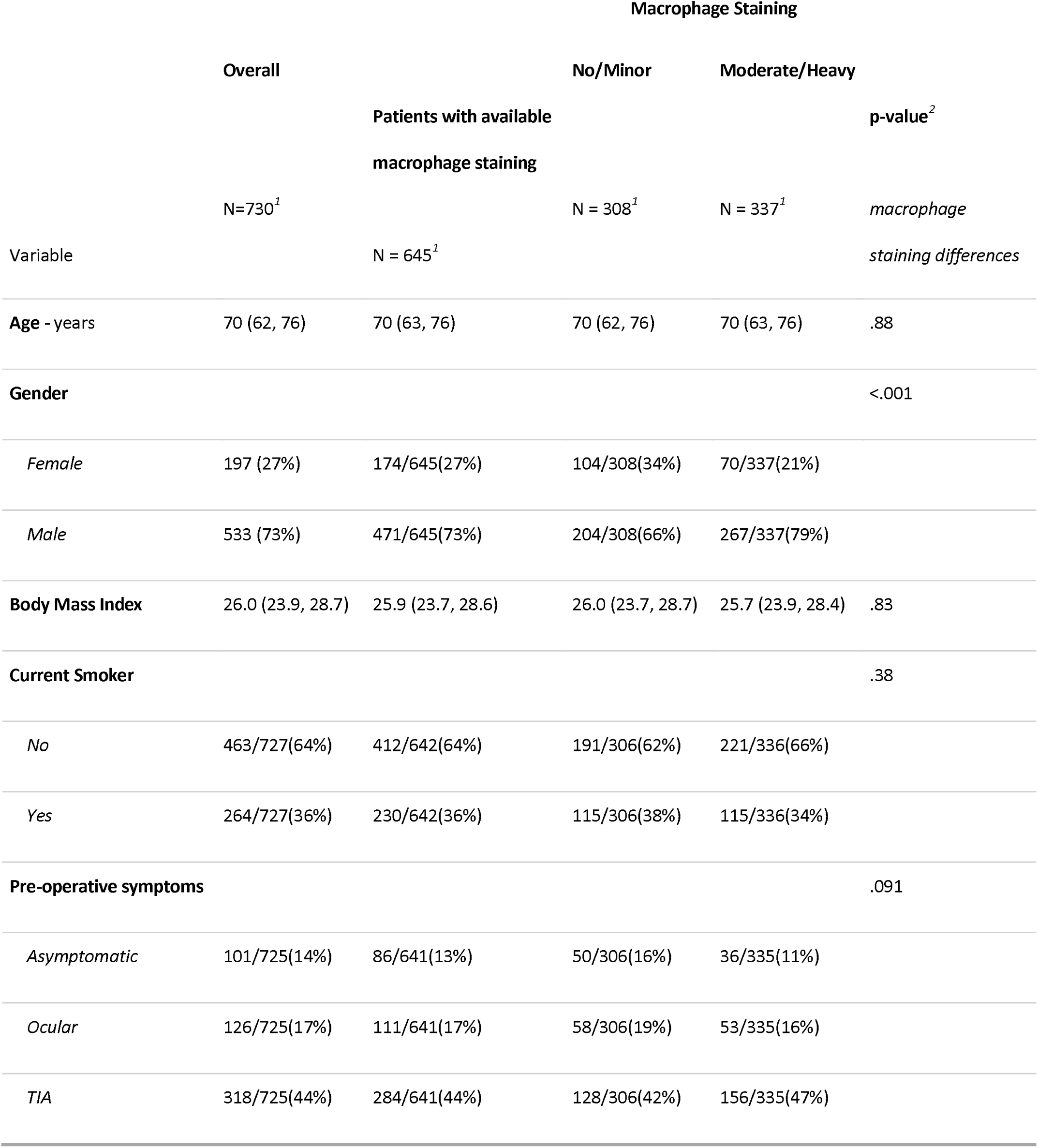
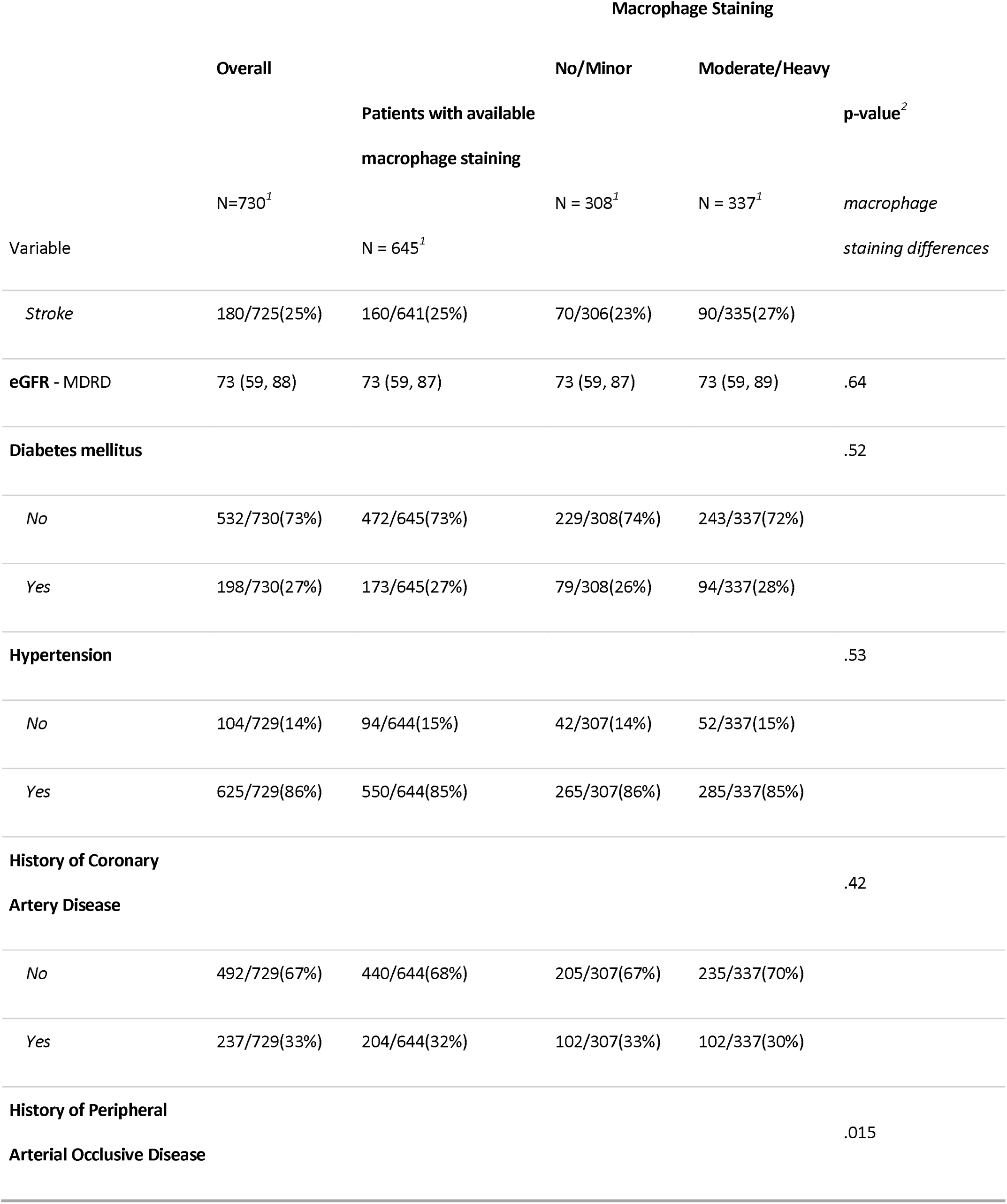
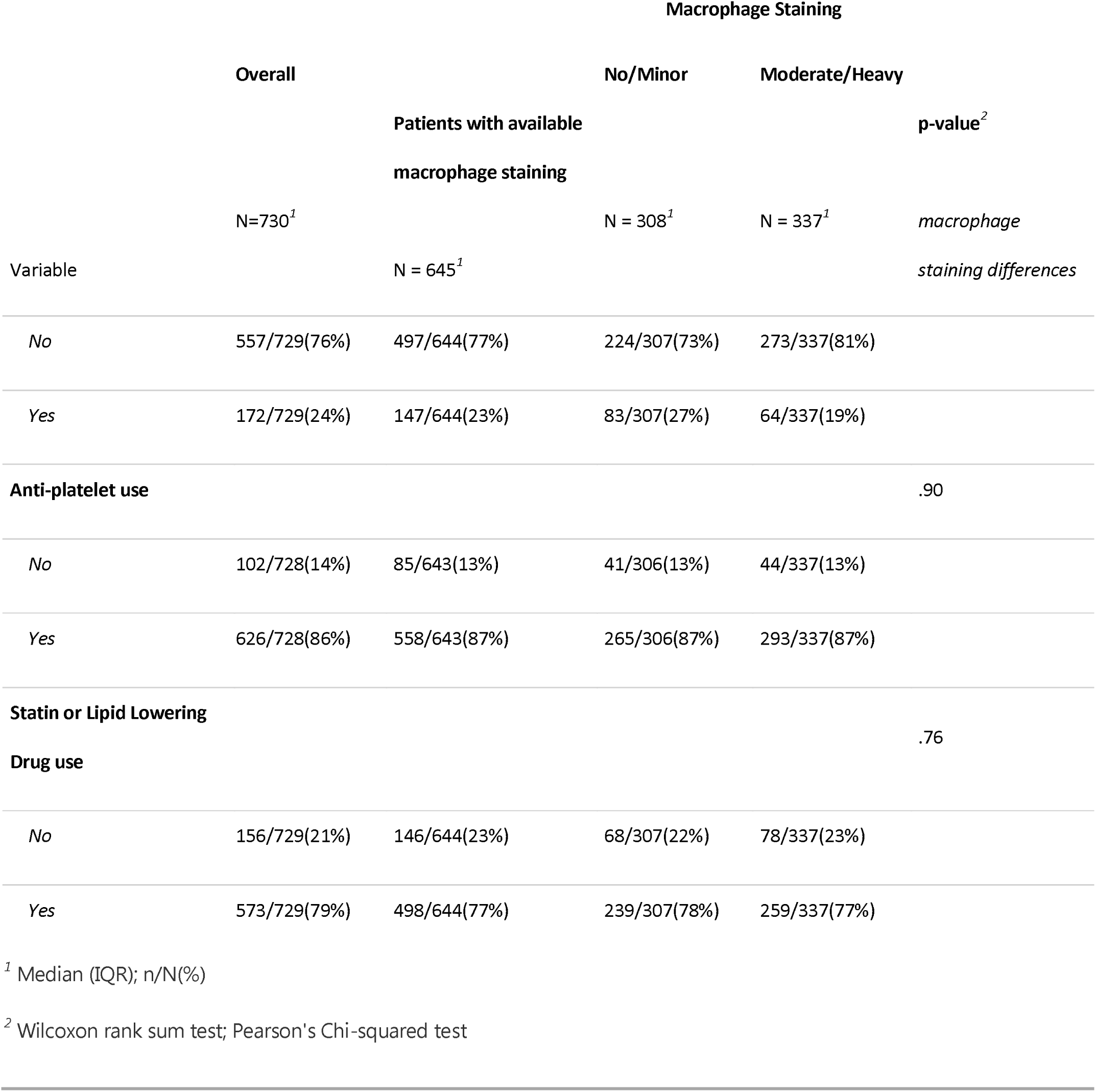
Baseline characteristics per macrophage staining group.

### Plasma levels of isoDGR-fibronectin and vulnerable plaque characteristics

Plasma levels of isoDGR-fibronectin were compared to 7 vulnerable carotid plaque characteristics that included macrophages (moderate/heavy staining), fat (>40% of plaque), presence of intraplaque hemorrhage, smooth muscle cells (no/minor staining), collagen (no/minor staining), calcification (moderate/heavy staining) and vessel density more or equal to median. Logistic regression analysis showed in both univariate (OR 1.39 (95% CI 1.06 – 1.83, p=.019)) and multivariate (OR 1.40 (95% CI 1.04 - 1.90, p=.028)) analysis higher odds ratio’s for moderate/heavy macrophage staining in plaques for patients with higher plasma levels of isoDGR-fibronectin. See table 2.

**Table 2.** Univariate and multivariate logistic regression analysis for plasma levels of isoDGR-fibronectin and plaque characteristics.

| Variable | Univariate |  | Multivariate |  |
| --- | --- | --- | --- | --- |
|  | OR (95% CI) | P value | OR (95% CI) | P value |
| Macrophages (moderate/heavy) | **1.39 (1.06 – 1.83)** | **0.019** | **1.40 (1.04 – 1.90)** | **.028** |
| Fat (>40%) | 1.32 (0.99 – 1.76) | 0.061 | 1.25 (0.91 – 1.73) | .17 |
| Intraplaque hemorrhage (present) | 1.17 (0.88 – 1.56) | 0.275 | 1.23 (0.91 – 1.68) | .85 |
| Smooth muscle cells (no/minor) | 0.89 (0.67 – 1.17) | 0.401 | 0.85 (0.63 – 1.15) | .30 |
| Collagen (no/minor) | 0.78 (0.57 – 1.08) | 0.140 | 0.80 (0.56 – 1.14) | .22 |
| Calcification (moderate/heavy) | 0.96 (0.74 – 1.26) | 0.787 | 1.01 (0.75 – 1.36) | .94 |
| Vessel density (≥ median) | 0.92 (0.66 – 1.27) | 0.598 | 0.98 (0.69 – 1.40) | .92 |

The multivariate logistic regression analysis is corrected for: age, sex, BMI, current smoking status, hypertension, diabetes, kidney function, statin or lipid lowering medication use, pre-operative symptoms, history of coronary artery disease and history of peripheral arterial occlusive disease.

### Plaque levels of isoDGR-fibronectin and adverse plaque characteristics

Since fibronectin is a matrix protein that is common in atherosclerotic plaques^19^, we measured isoDGR-fibronectin in plaques of all included patients for which frozen plaques were available (348 out of 730). Wilcoxon rank sum test showed higher median plaque levels of isoDGR-fibronectin in patients with moderate/heavy macrophage staining (0.043 (IQR 0.023 - 0.083) vs 0.033 (IQR 0.013 - 0.059), p=.005) when compared to the no/minor macrophage staining group. As for fat in plaques, median plaque levels of isoDGR-fibronectin were higher in plaques with fat percentages >40% (.051 (IQR 0.028 - 0.112) vs 0.034 (IQR) 0.014 - 0.056, p<.001) when compared to plaques with <40% fat. For intraplaque hemorrhage, median plaque isoDGR-fibronectin levels were higher in plaques showing intraplaque hemorrhage (0.032 (IQR 0.011 - 0.054) vs 0.043 (IQR 0.022 - 0.078), p=.005) when compared to plaques without intraplaque hemorrhage.

Comparison of smooth muscle cell staining showed a trend of higher median plaque levels of isoDGR-fibronectin with no/minor smooth muscle cells staining (0.044 (IQR 0.019 - 0.090) vs 0.036 (IQR 0.018 - 0.059), p=0.087). A similar trend was seen for higher median plaque levels of isoDGR-fibronectin in no/minor collagen staining groups, compared to the moderate/heavy collagen staining group (0.044 (IQR 0.023 - 0.072) vs 0.036 (IQR 0.018 - 0.065), p=.10). See figure 1 and table S1.

**Figure 1.**
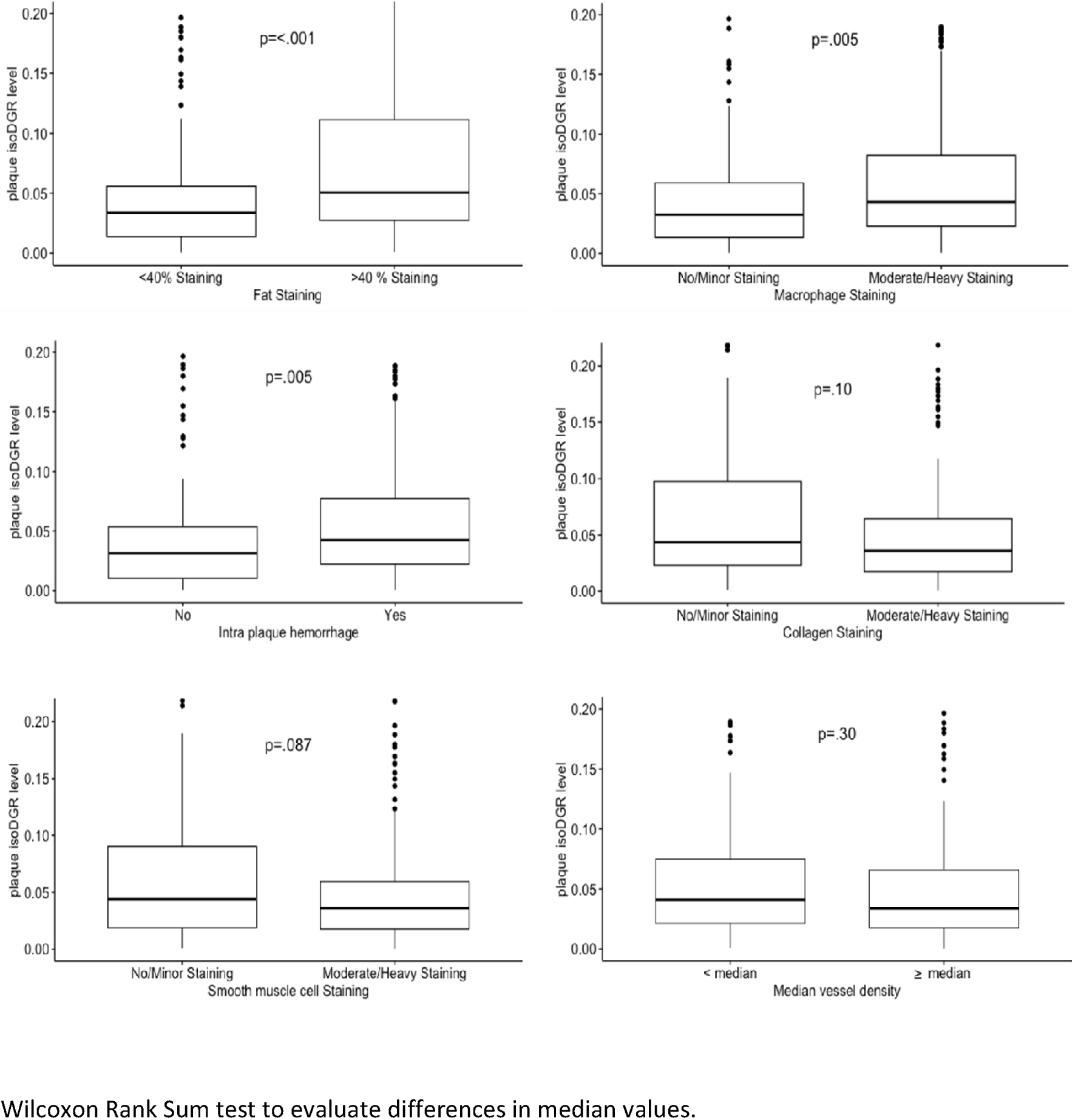
Boxplots of plaque levels of isoDGR-fibronectin in different plaque characteristics.

Logistic regression analysis showed that in both univariate and multivariate logistics regression analysis for adverse plaque characteristics, higher levels isoDGR-fibronectin result in a higher odds ratio’s for moderate/heavy macrophage staining (univariate OR 1.30 (95% CI 1.08 – 1.56), p=.006, multivariate OR 1.22 ( 95% 1.00 – 1.50), p=.049), >40% fat staining (univariate OR 1.51 (95% CI 1.22 – 1.90), p<.001, multivariate OR 1.44 (95% CI 1.14 – 1.86), p=.004) and presence of intraplaque hemorrhage (univariate OR 1.36 (95% CI 1.13 – 1.65), p=.001, multivariate OR 1.38 (95% 1.12 – 1.72), p=.003). See figure 2 and table S2.

**Figure 2.**
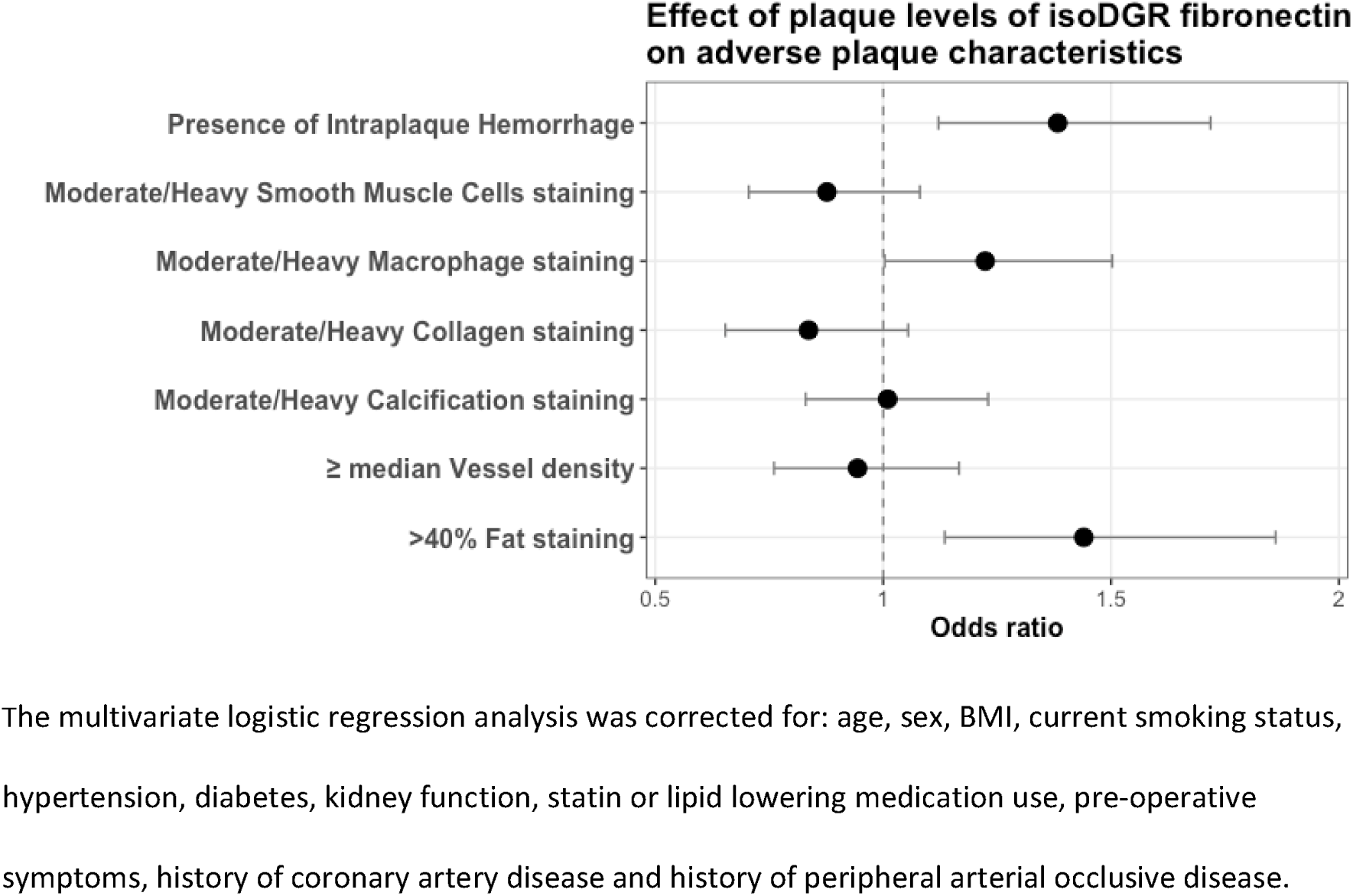
Forest plot for odds ratios in multivariate logistic regression analysis on plaque levels of isoDGR-fibronectin and adverse plaque characteristics.

### Plasma and plaque levels of isoDGR-fibronectin and plaque vulnerability score

Higher IsoDGR-fibronectin levels in plaque were positively associated with a higher plaque vulnerability score, with B coefficients of 0.27 (0.15 – 0,40, p<0.001) in the univariate model and 0.25 (0.12 – 0.39, p<0.001) in the multivariate model. There was no association between the plasma levels of isoDGR-fibronectin and the level of the plaque vulnerability score. See table 3.

**Table 3.** Univariate and multivariable adjusted beta coefficients of plasma and plaque levels of isoDGR-fibronectin in the linear regression model with the plaque vulnerability index.

| Variable | Univariate |  | Multivariate |  |
| --- | --- | --- | --- | --- |
|  | B coefficient (95% CI) | P value | B coefficient (95% CI) | P value |
| Plasma (continuous variable) | 0.16 (-0.05 - 0.36) | .13 | 0.17 (-0.04 - 0.39) | .12 |
| Plaque (continuous variable) | 0.27 (0.15 - 0.40) | **<.001** | 0.25 (0.12 - 0.39) | **<.001** |

## Discussion

IsoDGR-fibronectin has been shown to be involved in influx of macrophages in in vitro and mouse studies.^12,14,15^ In this study, we now show that high human plasma and plaque levels of isoDGR-fibronectin are not only associated with high atherosclerotic plaque macrophage levels but also with other adverse plaque characteristics in a large human cohort.

### Plasma isoDGR-fibronectin and plaque characteristics

First, high plasma levels of isoDGR-fibronectin were found in patients that had moderate/heavy macrophage staining in their plaque. This is in line with the previous study of Park et al.^12^, where isoDGR-fibronectin in plasma was measured in a cohort of 25 patients undergoing coronary artery bypass grafting (CABG) and compared to 25 controls lacking coronary artery disease (CAD). This study showed significant higher levels of plasma isoDGR-fibronectin in the patients with a high cardiovascular burden. Furthermore, this study showed that isoDGR-fibronectin activates pro-inflammatory pathways and integrins in both macrophages and endothelial cells, which are are key players in influx of macrophages.^20^ This is also in line with our study in PCMT1 ^-/-^ mice that reduced macrophage influx in body tissues when treated with a monoclonal antibody targeting isoDGR.^16^

### Plaque isoDGR-fibronectin and plaque characteristics

Next, IsoDGR levels in carotid plaques were analysed. Interestingly, higher plaque levels of isoDGR-fibronectin were not only associated with more macrophages, but also with a more atheromatous plaque and a higher likelihood of intraplaque hemorrhage. These results persisted when they were corrected for cardiovascular confounders. Next to this, a trend towards lower collagen and smooth muscle cell staining in the plaque was seen, contributing to a more vulnerable, rupture prone plaque.^21^

In previous studies, isoDGR-fibronectin has been detected in human atherosclerotic carotid plaques with proteomics.^22,23^ Dutta et al.^14^ showed in vitro that isoDGR-fibronectin leads to a higher monocyte/macrophage binding to isoDGR-fibronectin than to the normal fibronectin controls. The study of Kalailingam et al.^16^ further showed that isoDGR-fibronectin is involved in influx of macrophages and proinflammatory cytokines expression in several tissues. In atherosclerotic plaque formation, these macrophages contribute to foam cell formation^24^, which is in line with our finding of higher levels of atheroma in the plaque of patients with higher levels of isoDGR-fibronectin. Higher levels of macrophages and atheroma are both leading to higher local inflammation, resulting in neovascularization in the plaque. ^25^ This newly formed vasculature is often fragile and susceptible for leakage, leading to intraplaque hemorrhage^26^, which was also found to be associated with high isoDGR-fibronectin in this study.

In our study, plaque levels of isoDGR-fibronectin showed a higher correlation with adverse plaque characteristics than plasma levels of isoDGR-fibronectin. This could be attributed to the biology of fibronectin. Fibronectin is an extracellular matrix (ECM) protein essential in morphogenesis of vasculature and one of the earliest ECM proteins deposited on atherosclerotic prone sites, leading to atherosclerotic lesion formation^27,28^ and thus creating a very local effect with macrophage adhesion in isoDGR-fibronectin.^14^ Although plasma levels of fibronectin are higher in patients with cardiovascular disease, it was not able to predict the severity of disease in a study of Peng et al.^29^

### Plaque vulnerability score and plasma/plaque isoDGR-fibronectin

To quantify plaque vulnerability, a plaque vulnerability score was used, scoring one point for each individual vulnerable plaque characteristic. This showed a positive association of plaque levels of isoDGR-fibronectin and the plaque vulnerability score, both in the univariate and multivariate model. This result implies more vulnerable plaque structures when levels of isoDGR-fibronectin in plaque increases. There was no association with plasma levels of isoDGR-fibronectin and the plaque vulnerability score.

### Plaque isoDGR-fibronectin and MACE

Since plaque levels of isoDGR-fibronectin show higher associations with vulnerable plaque characteristics in our study, we looked for its association with MACE. Patients were dichotomized based on their plaque level of isoDGR-fibronectin above or below the overall median. MACE was found in 55 (15,8%) out of 348 patients with plaque samples available. Although MACE was more frequent in patients with isoDGR-fibronectin levels ≥ median (33 (19%) vs 22 (12,6%)), this reached no statistical significance in the Gray’s test, correcting for competing risk in the cumulative incidence function (p=.10). This could be due to a lack of power and that most local plaque characteristics are not associated with MACE.^7^

### Limitations

Some limitations of our study merit consideration. Unfortunately, only half of the included patients in our study had plaque samples available. For characteristics now only showing a trend, i.e. lower smooth muscle cell staining in plaques with higher levels of isoDGR-fibronectin or the association with MACE in the ≥ median plaque level of isoDGR group, a better significance might have been yielded with more plaque samples available. Nevertheless, associations with plaque levels of isoDGR-fibronectin and macrophages, fat and intraplaque hemorrhage are still robust. Secondly, the Athero-Express biobank does not contain information about the subtype of macrophages, or other plaque vulnerability indicators, such as B-cells, T-cells or mast cells. IsoDGR levels could therefore not be compared with more specific factors in atherosclerotic plaque inflammation. These results are found in a cohort of CEA patients and interpretation in other cohorts should be done with caution.

Despite these limitations, the significant strength of this study is that it is the first study to measure isoDGR-fibronectin in both plasma and plaque in a large human cohort of patients and associated with plaque characteristics.

### Importance

The importance of our study is that is shows for the first time that results from a large human cohort confirm previous in vitro and in vivo studies with murine models. This provides a better understanding of atherosclerotic plaque formation in humans. Moreover, it gives new insights in future perspective for potential therapies for atherosclerotic disease. In the study of Kalailingam et al^16^, Pcmt1 ^-/-^ mice, which cannot repair isoDGR motifs, were treated with either a monoclonal antibody targeting isoDGR or a placebo. The premature death observed in Pcmt1^-/-^ mice was significantly delayed in those treated with the monoclonal antibody. These mice not only showed lower levels of macrophages, but also had a lifespan three times longer than their placebo-treated counterparts.

## Conclusion

This study is the first study that shows an association with plasma and plaque levels of isoDGR-fibronectin and plaque levels of macrophages and other adverse plaque characteristics in a large human cohort. These findings attribute to a better understanding of plaque progression towards an adverse atherosclerotic plaque phenotype and identifies new potential treatment modalities.

## Data Availability

All data produced in the present study are available upon reasonable request to the authors

